# Infectious Complications Following CD30 Chimeric Antigen Receptor T-Cell Therapy in Adults

**DOI:** 10.1101/2024.07.10.24310235

**Authors:** Felicia Cao, Yueling Xiu, Michael Mohnasky, Jonathan S. Serody, Paul Armistead, Gianpietro Dotti, Melody Smith, Jonathan Huggins, Julia Messina, Bhanu Ramachandran, Jennifer Saullo, Joseph Stromberg, Manish K. Saha, Megan Walsh, Barbara Savoldo, Natalie Grover, Heather I. Henderson, Tessa M. Andermann

## Abstract

Infections are increasingly recognized as a common complication of chimeric antigen receptor (CAR) T-cell therapy. The incidence of clinically-defined infection after CD19.CAR T-cell therapy for relapsed/refractory lymphoma ranges from 60-90% in the first year after CAR T-cell therapy and is the most common cause for non-relapse mortality. However, infectious risk after CAR T-cell therapy targeting other malignancies is not well understood. Herein, we report for the first time, infectious complications after CD30.CAR T-cell treatment for patients with Hodgkin’s lymphoma and peripheral T-cell lymphoma. Since CD30 is only expressed on a subset of activated T and B-cells, we hypothesized that CD30.CAR T-cell patients would have reduced incidence and severity of infections after infusion compared to CD19.CAR T-cell patients. We retrospectively evaluated all 64 patients who received CD30.CAR T-cells at a single institution between 2016-2021, and assessed infections within one year after cell infusion, comparing these data to a contemporary cohort of 50 patients who received CD19.CAR T-cells at the same institution between 2018-2021. 23 CD30.CAR T-cell patients (36%) and 18 CD19.CAR T-cell patients (36%) developed a microbiologically confirmed infection. Infection severity and bacterial infections were higher in the CD19.CAR T-cell group compared to CD30.CAR T-cell recipients who more commonly had grade 1 respiratory viral infections. Our data reflect expected outcomes for severity and infection type in CD19.CAR T-cell patients and provide a benchmark for comparison with the novel CD30.CAR T-cell product. Although our findings require replication in a larger cohort, they have implications for antimicrobial prophylaxis guidelines after CD30.CAR T-cell therapy.

**KEY POINTS:**

1) The incidence of infections within the first year after CD30.CAR T-cell therapy was equivalent to that following CD19.CAR T-cell therapy
2) Viral infections were more common after CD30.CAR T-cell therapy but bacterial infections predominated after CD19.CAR T-cell therapy.

## INTRODUCTION

Chimeric antigen receptor (CAR) T-cell therapy has revolutionized the treatment of relapsed and refractory hematological malignancies. The first FDA approved CAR T-cells targeted CD19, a surface glycoprotein expressed on all B cells, for treatment of B cell malignancies including leukemia and lymphoma^1^. CD19.CAR T-cells are now the standard of care for patients with aggressive non-Hodgkin’s lymphoma who have progressed after two prior lines of therapy^2–6^.

Commonly recognized side effects of CAR T-cell treatment include cytokine release syndrome (CRS) and immune effector cell associated neurotoxicity (ICANS)^7,8^. However, infections are increasingly being recognized as common, and often severe, complications following CAR T-cell therapy. Infections are the most frequent cause of non-relapse related mortality after CD19.CAR T-cell therapy for patients with non-Hodgkin’s lymphoma^9,10^. The incidence of clinically-defined infections after CD19.CAR T-cell treatment ranged from 60 to 90% within one-year post-infusion, with 15-30% of infections being severe or life-threatening^9–12^. Infections were most commonly seen in the first month post CD19.CAR T-cell infusion, with the highest incidence of bacterial infections during this time period. Factors associated with infectious risk following CD19.CAR T-cell therapy included incidence of CRS/ICANS and corticosteroid use^9–11^. The CAR-HEMATOTOX model has been proposed to risk stratify patients treated with CD19.CAR T-cells who are at highest risk for hematotoxicity and severe infectious complications due to delayed hematopoietic recovery^13^.

In contrast, CD30 is transiently expressed by subsets of activated T and B cells and is highly expressed on hematological malignancies including Hodgkin’s lymphoma and anaplastic large cell lymphoma^14–16^. Brentuximab vedotin is a CD30 directed antibody-drug conjugate (ADC) that has been approved for treatment of patients with relapsed or refractory Hodgkin’s lymphoma^17,18^. Our group in a multi-center study characterized the outcome of patients, receivingCD30.CAR T-cell therapy and found an overall response rate (ORR) of 72% for the treatment of patients with relapsed/refractory Hodgkin lymphoma at 6 weeks post-treatment, with 36% progression-free survival over one year^19^. Another ongoing clinical trial is evaluating efficacy of CD30.CAR T-cells for patients with relapsed/refractory peripheral T-cell lymphoma (NCT04083495).

The patient population receiving CD30.CAR T-cell therapy differs substantially from CD19.CAR T-cell therapy, as Hodgkin’s lymphoma tends to affect younger patients who receive less cytotoxic chemotherapy and more ADC or immunotherapy based treatments^20,21^. Infectious risk after CD30.CAR T-cell therapy has not previously been characterized, including types of infections and their severity after CD30.CAR T-cell infusion. Currently, antimicrobial prophylaxis for any CAR T-cell treatment is based on existing guidelines for patients receiving CD19.CAR T-cells, although little data exists to inform prophylaxis after CAR T-cell treatments targeting different antigens^22,23^. Herein, we sought to characterize infectious risk during the first year after CD30.CAR T-cell infusion, benchmarked against infectious outcomes following CD19.CAR T-cell therapy, in order to more appropriately tailor antimicrobial prophylaxis for our patients.

## METHODS

### Patient cohort

Patients in this study were adults ≥18 years of age at the University of North Carolina at Chapel Hill (UNCCH) with relapsed or refractory CD30+ lymphomas as part of clinical trials NCT02690545, NCT03602157, NCT04083495, and NCT02663297. Of note, NCT02663297 investigated CD30.CAR T-cell infusion as consolidation after autologous stem cell transplant for CD30+ lymphoma (16 patients in this study)^24^. All patients who received CD30.CAR T-cell therapy from July 1, 2016 through July 31, 2021 were included in this retrospective review. For comparison, we included all adult patients ≥18 years of age at UNCCH who received CD19.CAR T-cells either as a commercial product or as part of a clinical trial (NCT03016377, NCT03696784, or NCT03594162) from April 1, 2018 through November 30, 2021. Consent was not required for inclusion in this study but was required for treatment with CD30.CAR T-cells. Approval was obtained from the UNCCH institutional review board (IRB#20-2745, PI: Andermann).

### Treatment and prophylaxis

Manufacture of CD30.CAR T-cells was performed as described previously^19^. Patients received 1-200 x 10^6^ CD30 targeted CAR-T cells for relapsed or refractory disease following lymphodepleting chemotherapy. Patients enrolled in NCT03602157 (12 patients) received CD30.CAR T-cells and additional T-cells co-expressing CD30.CAR and C-C Motif Chemokine Receptor 4 (CCR4). The majority of patients received antimicrobial prophylaxis consisting of valacyclovir 500mg daily or twice daily per provider preference and trimethoprim/sulfamethoxazole 160mg/800mg once per day, three times per week, starting at lymphodepletion (valacyclovir) or at day+30 (trimethoprim-sulfamethoxazole) until 6 months to one-year post-infusion or CD4 ≥200. Patients also received neutropenia prophylaxis with levofloxacin 500mg daily and fluconazole 400mg daily when absolute neutrophil count (ANC) ≤500 cells per mm2. The severity of CRS and ICANS were graded as described previously^25^. Patients with grade ≥2 CRS or ICANS were treated with tocilizumab (8mg/kg per dose IV) and/or corticosteroids as per institutional standards and previously published guidelines^25^.

### Data collection and infection definitions

Patient information was manually extracted from the electronic medical record and entered into a REDCap database by at least two independent reviewers (M.M., F.C., J.S., M.S., B.R.). Discrepancies between reviewers were resolved through adjudication by the primary study investigator (T.M.A.). Information regarding infections and antibiotics was abstracted starting 30 days prior through 365 days after CAR T-cell infusion. When censoring events were considered in select figures, patients were censored at relapse, next line of treatment, or at last contact with UNCCH.

Only microbiologically determined infections were included in this study, based on a positive culture or diagnostic test. One exception to this definition was the diagnosis of dermatomal Varicella Zoster Virus (VZV) which was included without an available positive diagnostic test if presentation met criteria as determined by the treating clinician. Bloodstream infections were adjudicated based on CDC/NHSN criteria^26^. Upper respiratory tract infections were based on clinical judgement and the absence of an oxygen requirement or infiltrates on chest imaging; lower respiratory tract infections were defined by infiltrates on chest imaging and lower respiratory tract symptoms. Infection severity was defined as mild (Grade 1), moderate (Grade 2), or severe (Grade 3) based on previously published Blood & Marrow Transplant Clinical Trials Network (BMT-CTN) 2023 criteria^27^.

### Statistical analysis

Continuous variables are reported as median and range; categorical variables are reported as number and percentage. For comparing characteristics between patients with and without infections, the Kruskal-Wallis test was used for continuous variables and Fisher’s exact test was used for categorical variables. The association between risk factors and time to infection among CD30.CAR T-cell patients was estimated using Fine-Gray subdistribution hazard models, treating all-cause mortality as a competing event^28^. ANC of 1.8 cells/mm^2^ was used as cut-off based on the institutional laboratory reference range. Age was modeled as a second-degree polynomial after evaluating its functional form in relation to infection risk. In cumulative-incidence curves, patients contributed time from the date of infusion until the earliest of the following: first infection (overall and stratified by pathogen type), death, or administrative censoring at 365 days post-infusion. As a secondary analysis, patients were censored if relapse occurred within one year after infusion. P-values less than 0.05 were considered statistically significant. Analyses were performed using R software version 4.3.2, with the *cmprsk* package used to estimate the cumulative incidence of infection.

## RESULTS

### Patient characteristics

Sixty-four adult patients received CD30.CAR T-cells at the University of North Carolina at Chapel Hill (UNCCH) from 2016 to 2021 (Table 1). Fifty patients (78%) had Hodgkin’s lymphoma (HL) and 14 patients (22%) had peripheral T-cell lymphoma. Patient characteristics for CD30.CAR T-cell treated patients are described in **Table 1**, stratified by infection status. The median age was 40.9 (range 18 to 77) years. This was a heavily pre-treated patient population, with patients receiving a median of four lines of therapy (range 2 to 17) prior to CAR T-cell therapy. Fifty-six patients (88%) had received a hematopoietic stem cell transplantation (HSCT) prior to CAR T-cell treatment, with the vast majority being autologous transplants.

**Table 1.**
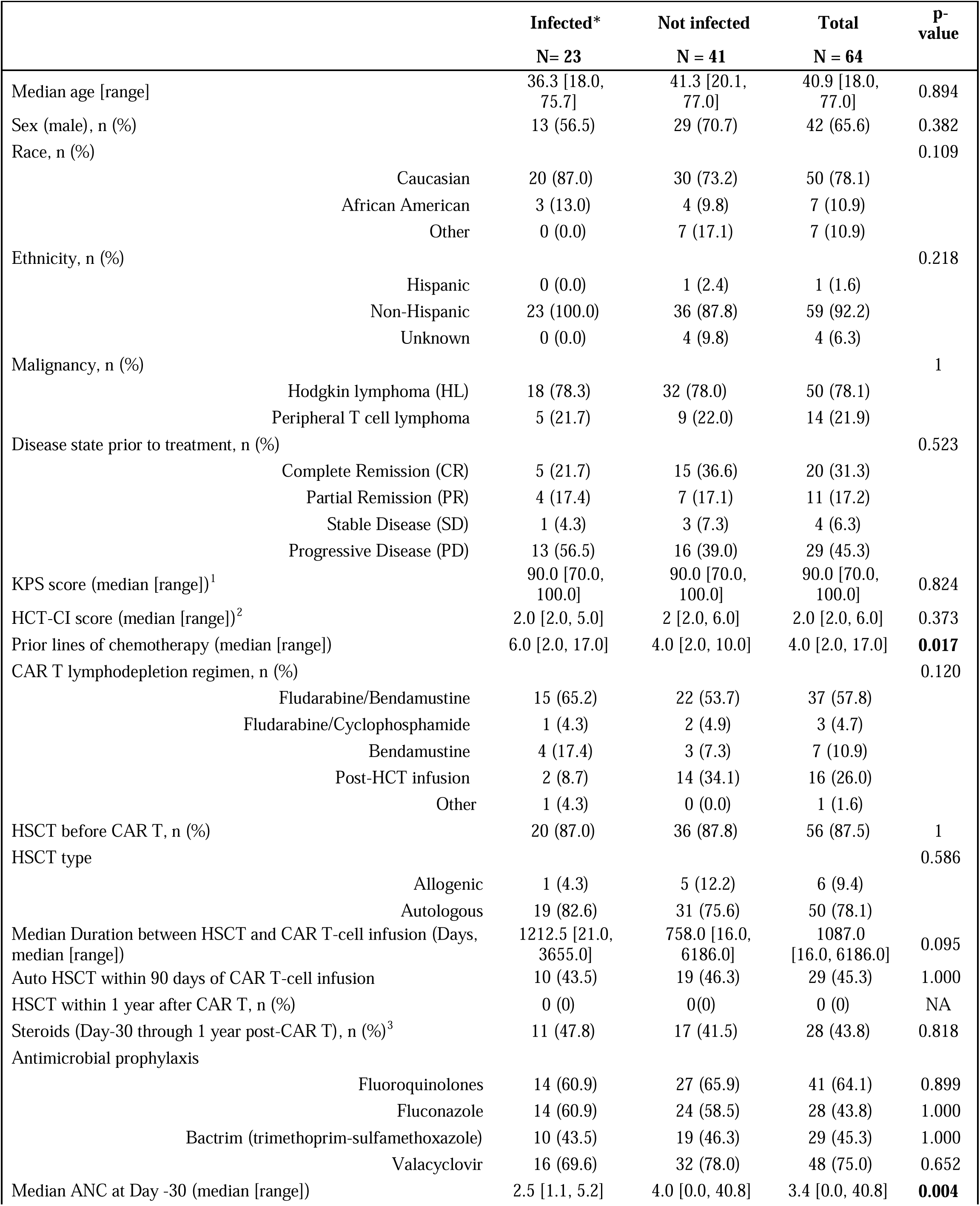
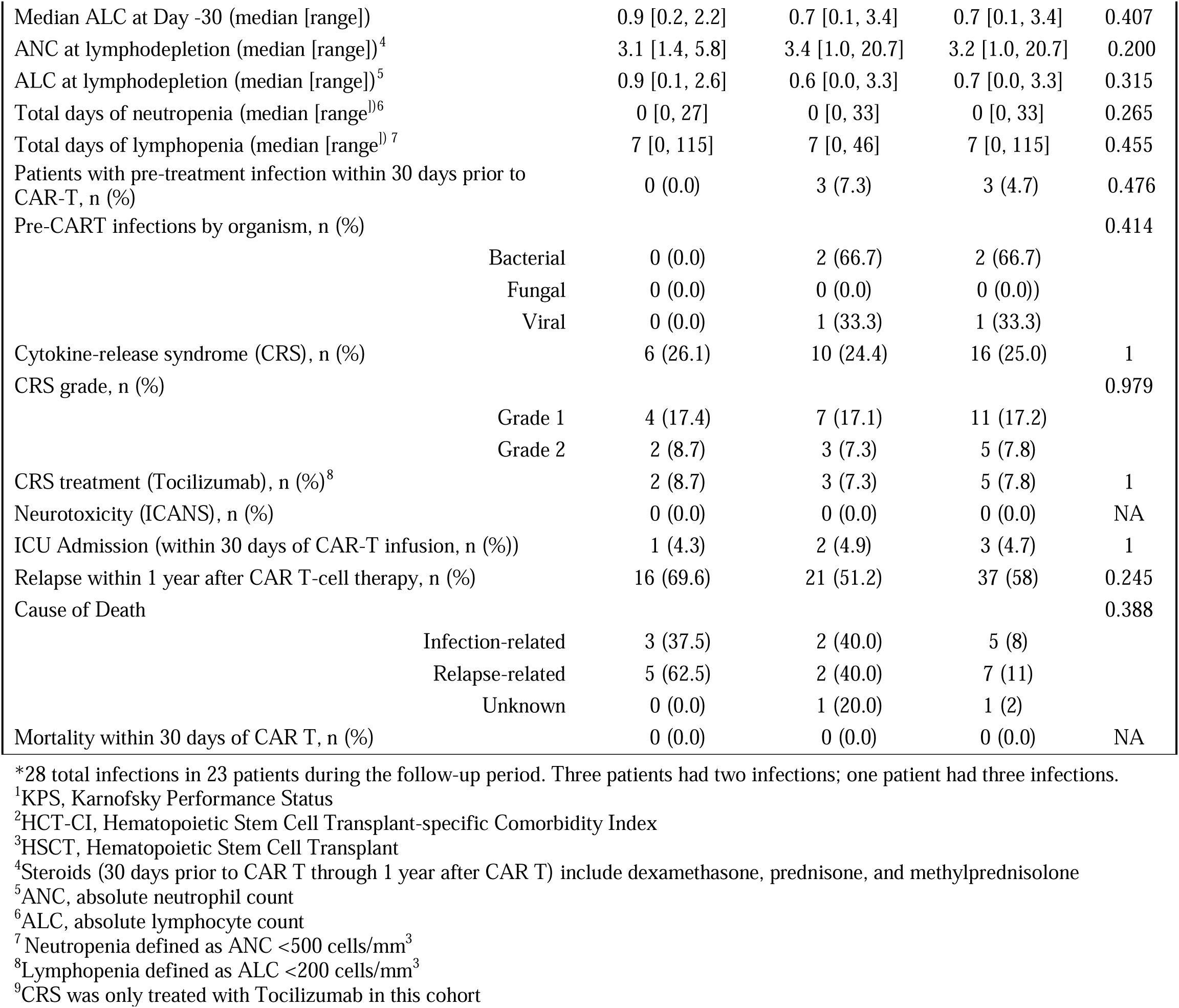
Demographics and treatment-related complications of patients with and without microbiologically confirmed infections within one year after anti-CD30 CAR-T therapy.

For comparison, we included the cohort of all 50 patients treated with CD19.CAR T-cells at UNCCH from 2018 to 2021 (**Supplementary Table S1)**. Twelve patients within this group (24%) had acute lymphoblastic leukemia (ALL), 30 (60%) had diffuse large B-cell lymphoma (DLBCL), and eight (16%) had follicular lymphoma or mantle cell lymphoma. The median age was 60.6 (range 21 to 81) years. Similar to the CD30.CAR T-cell therapy cohort, patients were heavily pre-treated, having received a median of four (range 2 to 12) prior treatment lines. The proportion of those receiving CD19.CAR T-cells who had received a prior HSCT (14 patients, 28%) was lower compared to those receiving a CD30 product at our institution.

The incidence of cytokine release syndrome (CRS) and immune effector cell-associated neurotoxicity syndrome (ICANS) was lower in the CD30.CAR T-cell cohort (**Table 1**) compared to the CD19.CAR T-cell cohort (**Supplementary Table S1)**. Sixteen patients (25%) who received CD30.CAR T-cells experienced CRS of any grade, with only five patients (8%) demonstrating Grade 2 CRS, and none with Grade 3 or higher CRS. No patients were diagnosed with ICANS after CD30.CAR T-cell infusion. In contrast, 34 patients (68%) in the CD19.CAR T-cell group developed CRS of any grade, with 16 patients (32%) who developed Grade 2 CRS and one patient (2%) who developed Grade 3 CRS. 14 patients (28%) developed ICANS after CD19.CAR T-cell therapy.

### Incidence of infections after CAR T-cell therapy

Twenty-three patients (36%) developed a microbiologically confirmed infection within one year after CD30.CAR T-cell therapy. The cumulative incidence of all infections within one year after CD30.CAR T-cell treatment is shown in **Figure 1**. Six bacterial infections, 21 viral infections, and one fungal infection were seen in 23 CD30.CAR T-cell patients. When censoring for relapse, three bacterial infections and 16 viral infections were reported in 16 CD30.CAR T-cell patients (**Supplementary Figure S1**). No fungal infections were observed after censoring for relapse.

**Figure 1.**
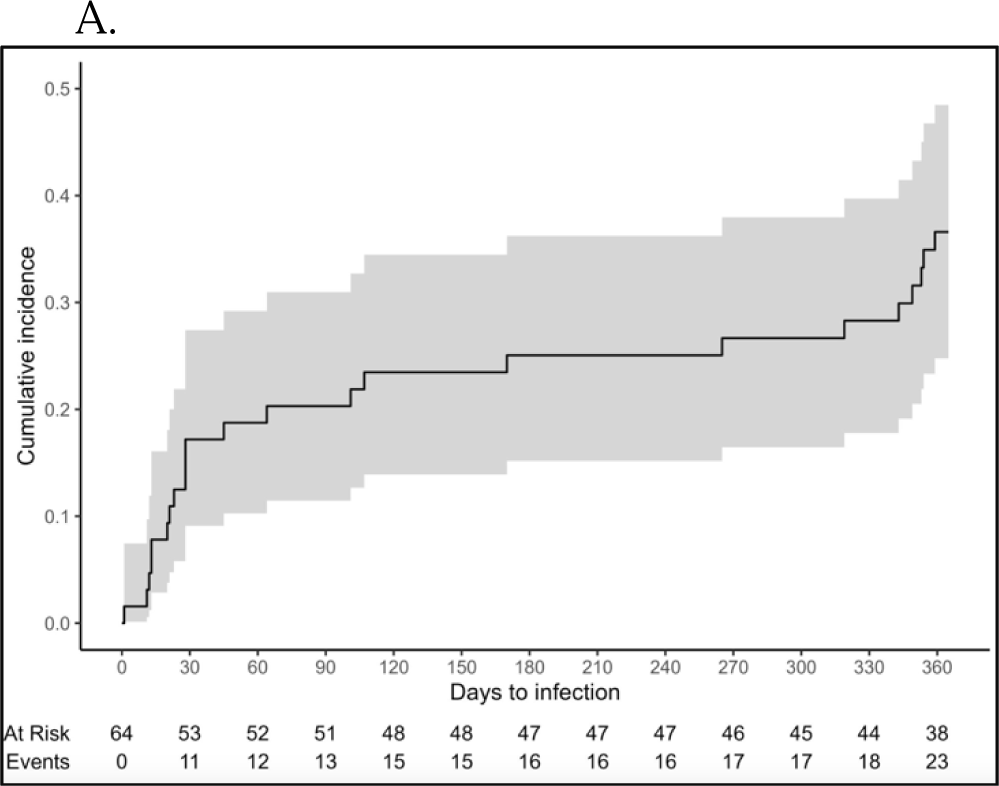
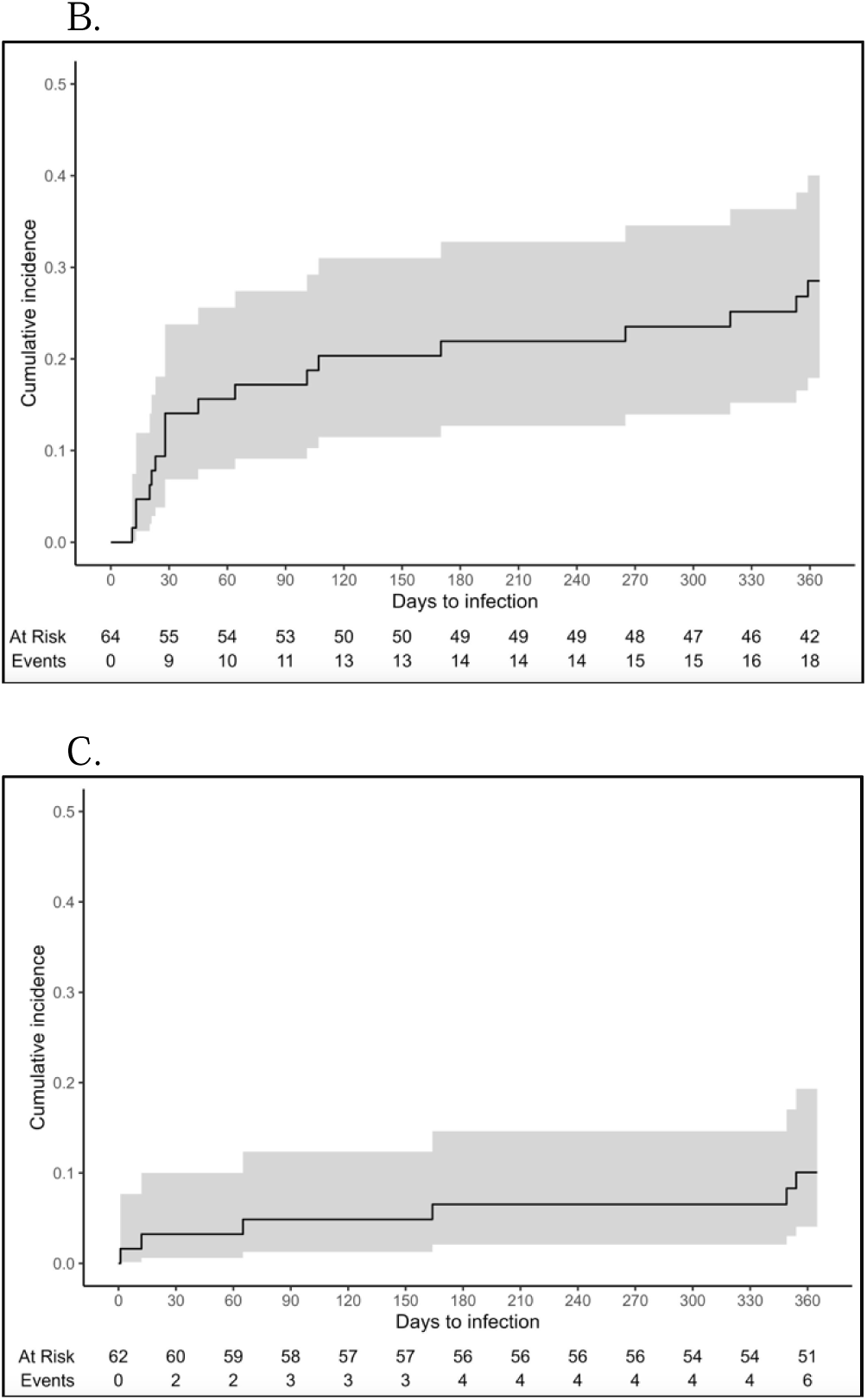
Greater numbers of viral infections were observed after CD30. CAR T-cell therapy compared to bacterial infections. Cumulative incidence curves demonstrating time to first infections in the one year after CD30 CAR T-cell therapy, including all infections (A), only viral infections (B), and only bacterial infections (C).

In comparison, the same proportion of CD19.CAR T-cell patients (n=18, 36%) developed an infection within one year from cell infusion (**Figure 1**). Eighteen bacterial infections, 12 viral infections, and three fungal infections were reported in 18 infected CD19.CAR T-cell patients. When censoring for relapse, seven bacterial infections and six viral infections were reported in 10 patients (**Supplementary Figure S1**).

The incidence of bacterial, viral, and fungal infections from days 0-28, days 29-90, and days 91-365 for CD30 and CD19.CAR T-cell patients are shown in **Figure 2**. In the CD30.CAR T-cell cohort, viral infections were more common than bacterial infections at all timepoints. This contrasts with CD19.CAR T-cell patients, in which bacterial infections were more common than viral infections, most prominently in the first 28 days after CAR T-cell infusion. This difference holds even when censoring for relapse (**Supplementary Figure S1**). Fungal infections were rare, occurring only in one patient who received CD30.CAR T-cells and three patients who received CD19.CAR T-cells, and only in the setting of relapsed disease. A list of causative infectious organisms is shown in **Table 2**, which again highlights that viral infections were more common after CD30.CAR T-cell therapy, and bacterial infections are more common after CD19.CAR T-cell treatment.

**Figure 2.**
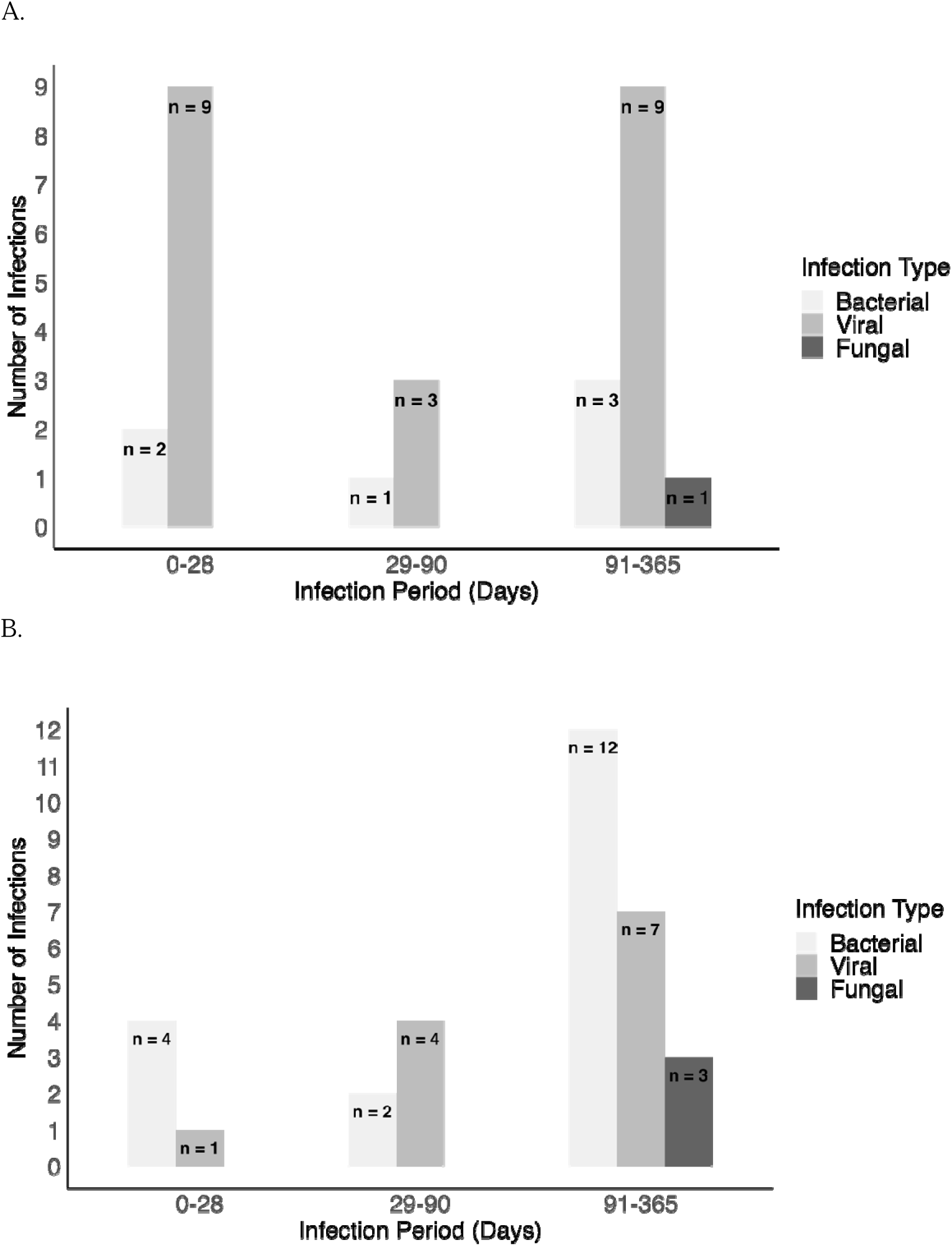
Similar numbers of microbiologically confirmed infections were observed between CD30 and CD19.CAR T-cell recipients. Number of infections including bacterial, viral, and fungal etiologies during the first 1 year following CAR T-cell therapy, broken into infection periods (0-28, 29-90, and 91-365 days) and compared between anti-CD30 CAR-T therapy (A) and anti-CD19 CAR-T therapy (B). All available data are shown, regardless of relapse.

**Table 2.** Infectious organisms subdivided by bacterial, viral, or fungal etiology after (A) CD30 CAR T-cell therapy or (B) CD19 CAR T-cell therapy. Number in parathesis indicates total number of infections for causative organism. EBV = Epstein Barr Virus. HSV = Herpes Simplex Virus. RSV = Respiratory Syncytial Virus. SARS-CoV-2 = COVID-19. VZV = Varicella Zoster Virus.

| Infections after CD30 CAR T-cell therapy |  |  |
| --- | --- | --- |
| Bacterial | Viral | Fungal |
| *Escherichia coli* | Coronavirus NL63 | *Pneumocystis jirovecii* |
| *Serratia marcescens* | EBV (2) |  |
| *Staphylococcus aureus* (2) | HSV |  |
| *Staphylococcus epidermidis* | Influenza A |  |
| *Streptococcus dysgalactiae* | Rhinovirus (6) |  |
|  | RSV (2) |  |
|  | SARS-CoV-2 (6) |  |
|  | VZV (5) |  |

B.
| Infections after CD19 CAR T-cell therapy |  |  |
| --- | --- | --- |
| Bacterial | Viral | Fungal |
| Bacterial vaginosis | HSV (3) | *Fusarium species* |
| *Campylobacter spp.* | Norovirus | *Pneumocystis jirovecii* (2) |
| *Citrobacter braakii* | Rhinovirus (3) |  |
| *Enterococcus faecalis* | RSV |  |
| *Escherichia coli* | SARS-CoV-2 (4) |  |
| *Klebsiella pneumoniae* (3) |  |  |
| *Staphylococcus aureus* |  |  |
| *Serratia marcescens* |  |  |
| *Staphylococcus epidermidis* |  |  |
| *Streptococcus agalactiae* |  |  |
| *Streptococcus mitis* |  |  |

Notably, the incidence of infections was higher in patients with relapsed disease after day 90, as 13 infections were seen during this time in the CD30.CAR T-cell cohort and 22 infections in the CD19.CAR T-cell cohort. However, this appears to correlate with disease recurrence as there were only five infections that occurred in the CD30.CAR T-cell cohort and three infections in the CD19.CAR T-cell cohort in non-relapsed patients. Nonetheless, similar to previously published data for CD19.CAR T-cell therapy, patients receiving CD30.CAR T-cell therapy may be at increased risk for infectious complication for a prolonged period post-cell infusion, although bacterial infections were more common in the first 28 days in both our data and prior literature^9–12^.

### Severity of infections after CAR T-cell therapy

To further understand the clinical relevance of infections seen after CAR T-cell therapy, the severity of infections was assessed using BMT-CTN 2023 criteria^27^. The severity of microbiologically confirmed infections within one year after CD30 or CD19.CAR T-cell infusion is shown in **Figure 3**. Similar data, censored for relapsed disease, are shown in **Supplementary Figure S2**. For CD30.CAR T-cell patients, most infections were mild with three grade 1 bacterial infections and 18 grade 1 viral infections. Severe or life-threatening infections were rare, with only one grade 3 bacterial infection and one grade 3 viral infection found in the CD30.CAR T-cell cohort. When censoring for relapse within one year of CD30.CAR T-cell infusion, only one grade 3 viral infection (of a total of 19 infections) remained.

**Figure 3.**
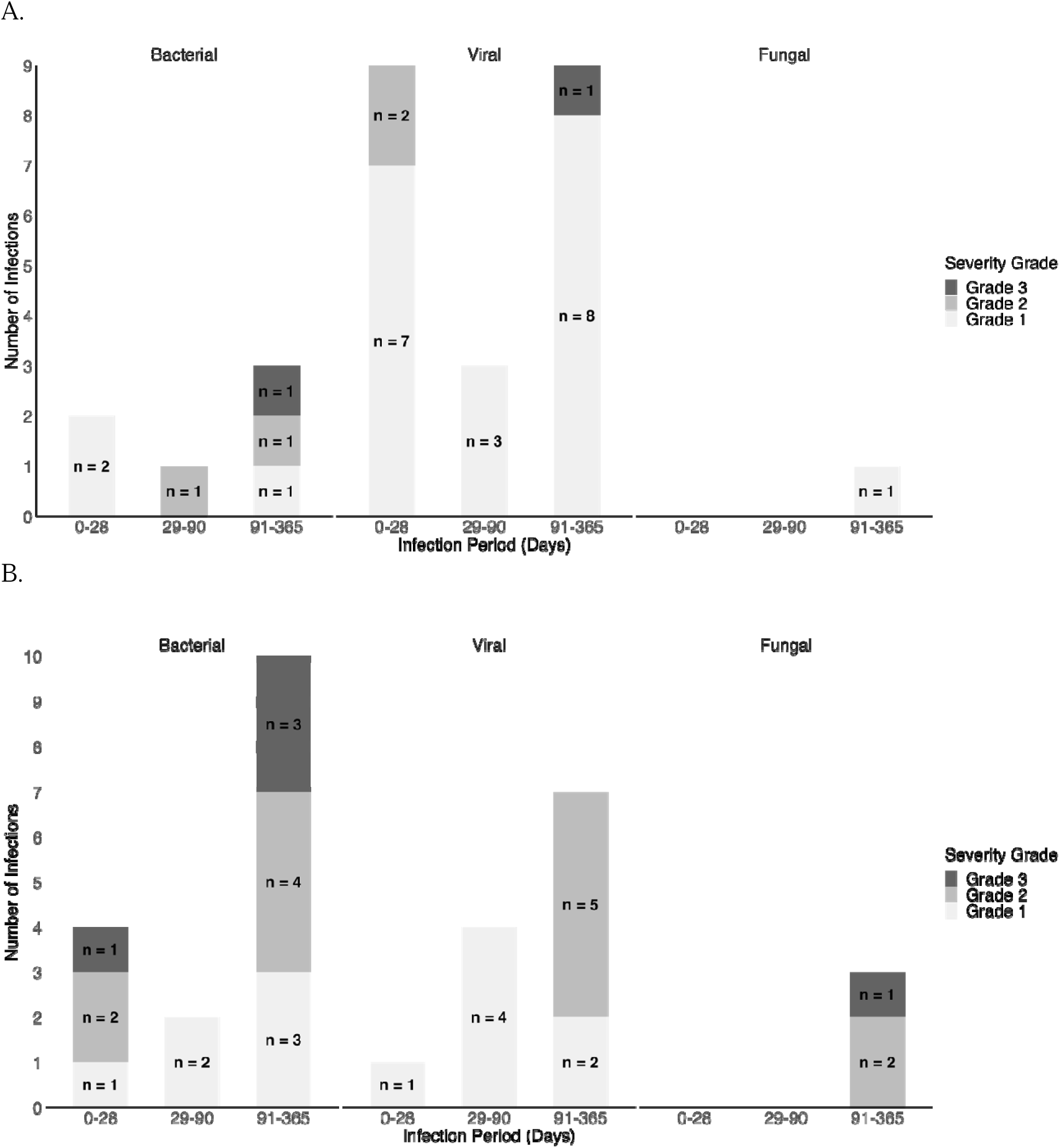
More severe infections and more infections of bacterial origin were observed after CD19.CAR T-cell therapy compared to CD30.CAR T-cell therapy. Number of infections including bacterial, viral, and fungal etiologies of different severities occurring during the first 1 year following CAR T-cell therapy, broken into infection periods (0-28, 29-90, and 91-365 days), and compared between anti-CD30 CAR-T therapy (A) and anti-CD19 CAR-T therapy (B). Of note, two infections in B are not shown as their severity was determined as “unknown”. All other available data are shown, regardless of relapse.

In contrast, the CD19.CAR T-cell patients had more severe infections overall with four grade 3 bacterial infections but no severe or life-threatening viral infections. When censoring for relapse, there were two grade 3 bacterial infections (total n=13 infections) within one year of CD19.CAR T-cell infusion. These data demonstrate that overall severity of infection was lower in the CD30.CAR T-cell patients compared to the CD19.CAR T-cell cohort. In summary, most infections in the CD30.CAR T-cell group were caused by respiratory viral pathogens and were milder whereas more patients in the CD19.CAR T-cell group had bacterial infections with higher severity.

### Factors associated with increased infectious risk

Baseline characteristics of the 23 patients who developed a microbiologically confirmed infection in the first year after CD30.CAR T-cell therapy were compared to the 41 patients who did not develop an infection in **Table 1**. In univariate analysis, the only baseline characteristic significantly different between these two groups was ANC at 30 days prior to cell infusion (p=0.004). After adjusting for age, ANC ≤1.8 cells/mm^2^ at day −30 remained significantly associated with infection risk, with an adjusted hazard ratio (HR) of 3.4 (95% confidence interval [CI] 1.5 – 8.1). The small number of patients in our study limited our ability to include additional covariates in our model.

Steroid use was not associated with increased infection risk in CD30.CAR T-cell patients, with 11 patients (48%) receiving steroids within a year of cell infusion in the infected group and 17 patients (42%) receiving steroids within a year of cell infusion in the non-infected group. Steroid use was generally low dose (less than or equivalent to 40mg prednisone daily) or a one-time single dose in the CD30.CAR T-cell group; in this cohort, steroids were not used for treatment of CRS/ICANS, and instead were given for a variety of indications including premedication prior to chemotherapy or surgery, or for adrenal insufficiency. This contrasts with CD19.CAR T-cell patients, in which steroid use was mostly for treatment of CRS/ICANS and significantly associated with infection risk (p=0.02) (**Supplementary Table S1**). Steroid use was higher in the infected group, likely due to treatment for ICANS (n=9, 50%) compared to patients who had no infections (n=5, 15.6%, p=0.02 for infected v. non-infected). Notably, this same relationship between steroid use and infectious risk has been observed in prior studies in CD19.CAR T-cell recipients^10,11^. Infection-related mortality was a significant cause of death in the CD19.CAR T-cell group, with 7 (63.6%) infection-related deaths and only 4 (36.5%) relapsed-related deaths in the infected group compared to 13 (86.7%) relapse-related deaths and 2 (13.3%) “other” causes of death in the non-infected group (p=0.004). Overall, these data show that CD19.CAR T-cell patients have more measured variables associated with infection risk in univariate analysis compared to CD30.CAR T-cell patients.

### Immune reconstitution after CD30.CAR T-cell therapy

Given the significant association between day-30 ANC and infectious complications in CD30.CAR T patients, we sought to better understand the trajectory of immune reconstitution after CD30.CAR T-cell therapy. We examined the kinetics of absolute neutrophil and lymphocyte counts (ANC and ALC) 30 days prior to, and at multiple timepoints after, CAR T-cell infusion. Differences in ANC and ALC in CD30.CAR T-cell patients with and without infections are shown in **Figure 4**. The nadir for ALC was on day 0 of CAR T-cell infusion as expected, secondary to lymphodepleting chemotherapy. Surprisingly, the nadir for ANC was seen on day 60, a finding which cannot be explained by lymphodepletion. There were no significant differences in ANC and ALC between the infected and non-infected groups over time except for a lower ANC 30 days prior (p=0.004) and at day 0 (p=0.01) in those receiving CD30.CAR T-cells who were infected. Similarly, when comparing ANC and ALC for infected and non-infected CD19.CAR T-cell patients, there were no significant differences in ANC and ALC at most timepoints. The only exceptions were for ANC at 9 months post-cell infusion (p=0.02) and ALC at 30 days prior to cell infusion (p=0.04); in both instances, cell counts were higher in infected patients, a finding likely spurious due to multiple testing.

**Figure 4.**
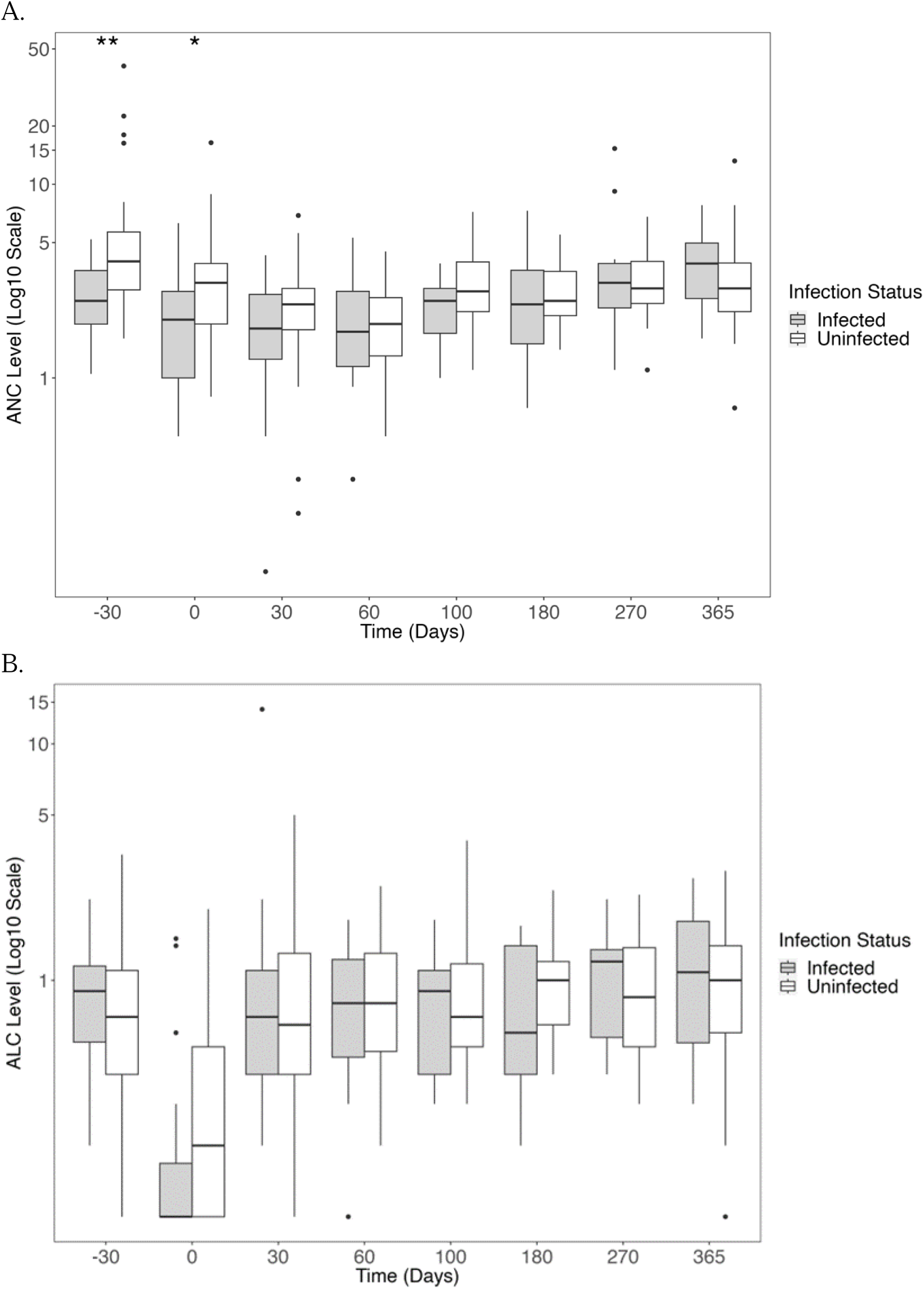
Immune recovery after CD30 CAR T-cell therapy demonstrates rare differences between infected and uninfected patients within 1 year after infusion. Absolute neutrophil counts (ANC, Figure 4A) and absolute lymphocyte counts (ALC; Figure 4B) are demonstrated at timepoints relative to the day of CAR-T infusion. Com arisons between infected and uninfected patients were performed using the Wilcoxon rank-sum test, with p-value <0.05 being given one star and p-value <0.01 being given two stars at the top of the plot.

There were significant differences seen when comparing ANC and ALC between CD30 and CD19.CAR T-cell patients as seen in **Figure 5**. There was no difference in ANC and ALC 30 days prior to cell infusion in the two cohorts. However, significant differences were seen at multiple timepoints on and after cell infusion. ANC was significantly lower in the CD19.CAR T-cell cohort compared to the CD30.CAR T-cell group at every timepoint on or after cell infusion except at day 60 and 1 year. Similar results were seen for ALC, which was significantly lower in the CD19.CAR T-cell versus CD30.CAR T-cell cohort at every timepoint on or after cell infusion except for day +60. Of note, these statistical comparisons were performed after removing CD30.CAR T-cell patients enrolled in clinical trial NCT02663297, since an immediately preceding autologous HSCT may have influenced blood counts^24^. However, since the statistical trends were similar, all patient data was shown in the figure.

**Figure 5.**
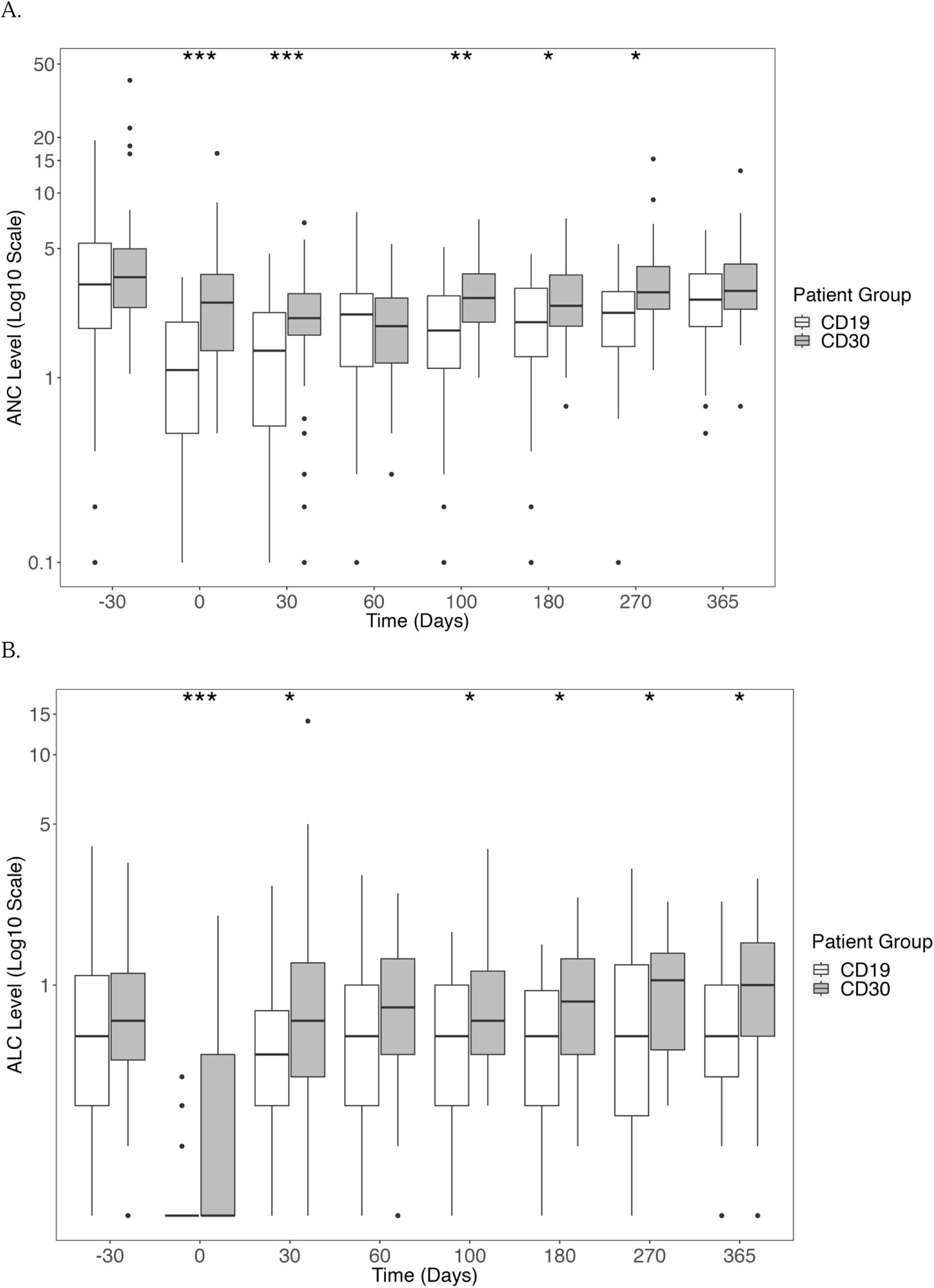
Immune recovery after CAR T-cell therapy demonstrates significant differences between CD30 and CD19 products within 1 year after infusion. Absolute neutrophil counts (ANC, Figure 5A) and absolute lymphocyte counts (ALC; Figure 4B) are demonstrated at timepoints relative to the day of CAR-T infusion. Comparisons between CD30 and CD19 recipients were performed using the Wilcoxon rank-sum test, with p-value <0.05 as one star, p-value <0.01 with two stars, and p-value <0.005 with three stars at the top of the plot.

## DISCUSSION

In these two cohorts of 114 patients in total treated with CD30 or CD19.CAR T-cells for relapsed/refractory hematological malignancies, we found a similar incidence of microbiologically confirmed infections within one year from cell infusion. Notably, when censoring for relapse, the incidence of infection was higher in CD30.CAR T-cell (27%) versus CD19.CAR T-cell recipients (16%). Infections in the CD30.CAR T-cell cohort were primarily viral (versus bacterial in the CD19.CAR T-cell group) and were less severe. As expected, for both groups, a greater proportion of infections were seen in the initial 30-day period after cell infusion. Unexpectedly, the higher incidence of viral infections in the CD30.CAR T-cell cohort is not related to lymphopenia, as ALC was significantly lower at most timepoints after CD19.CAR T-cell infusion compared to CD30.CAR T-cell therapy. Instead, ANC 30 days prior to CAR T infusion was significantly lower in infected patients.

There were multiple differences between the two cohorts which may contribute to variability in infection types and severity. Yet, the CD19.CAR T-cell treated cohort remains an appropriate benchmark for comparison in our review of infection outcomes following CD30.CAR T-cell treatment, with results from our CD19 cohort similar to those published previously.^9–12^ One important difference between the two cohorts is the inclusion of a majority of patients with Hodgkin’s lymphoma in the CD30 group, as these patients tend to be younger at diagnosis. This was reflected in the lower median age for the CD30 compared to the CD19 cohort (40.9 v. 60.6 years). Additionally, most patients in the CD30 cohort had received prior HSCT, including 29 total patients who received autologous HSCT within 90 days prior to CD30.CAR T-cell therapy. Despite this additional immunosuppression in the CD30 group, infection risk was much less frequent and less severe.

Antimicrobial prophylaxis use, including both antibacterials and antivirals, was similar between infected and non-infected patients in both the CD30 and CD19.CAR T-cell cohorts, but overall higher in the CD19.CAR T-cell treated patients (**Tables 1 and Supplementary Table S1**). We postulate this to be a consequence of more standardized and increased antimicrobial prophylaxis use in patients post CD19.CAR T-cell therapy (i.e., patients underwent CD30.CAR T-cell treatment starting 2 years earlier (2016 v. 2018) than CD19.CAR T-cell patients). With regards to antiviral prophylaxis, five cases of VZV infection were observed in CD30.CAR T-cell patients while none were in CD19.CAR T-cell patients. This may be related to a higher proportion of patients on valacyclovir prophylaxis in the CD19 cohort as four of the five patients who developed VZV reactivation were not on prophylaxis at time of infection.

Another difference between the two cohorts was the incidence of CRS/ICANS, which was far less frequent and less severe in CD30.CAR T-cell patients. No patients developed ICANS after CD30.CAR T-cell infusion compared to 28% of CD19.CAR T-cell patients. The difference in ICANS/CRS between the two cohorts could be an explanation for why steroid use was significantly linked with infections (p=0.02) in the CD19.CAR T-cell cohort but not in the CD30.CAR T-cell cohort (p=0.8) as overall high-dose steroid use was much more common in the former (**Table 1 and Supplementary Table S1**).

Another complicating factor may be the differences in lymphodepletion regimens between the two cohorts. The majority of CD30.CAR T-cell treated patients received fludarabine and bendamustine for lymphodepletion (n=37, 57.8%) with other regimens consisting of fludarabine/cyclophosphamide (n=3, 4.7%), bendamustine (n=7, 10.9%), and autologous HSCT (n=16, 26.0%). In contrast, the majority of CD19.CAR T-cell receiving patients were treated with fludarabine/cyclophosphamide (n=44, 88%) with some patients receiving fludarabine/bendamustine (n=5, 10%) or bendamustine alone (n=1, 2%) as alternative lymphodepletion regimens (**Table 1 and Supplementary Table S1**). Bendamustine use prior to apheresis for CD19.CAR T-cell therapy has been associated with lower T-cell counts at apheresis.^29^. However, since ALC at multiple timepoints post infusion was lower in the CD19.CAR T-cell cohort, who mostly did not receive bendamustine compared to CD30.CAR T-cell cohort, it seems unlikely that lymphodepletion alone is enough to account for differences in infections and immune recovery.

Strengths of this study include a relatively large total cohort of 114 CAR T-cell recipients and inclusion of only microbiologically confirmed infections, with far less between-clinician bias. To our knowledge, this is the first report of infection incidence and severity for patients treated with CD30.CAR T-cell therapy, broadening our understanding of the epidemiology of infectious risk post-CAR T-cell infusion. Limitations include heterogeneous disease types that preclude direct comparison between cohorts, with more than four different hematological malignancies represented (Hodgkin’s lymphoma, DLBCL, ALL, and peripheral T cell lymphoma). Lymphocyte subsets and immunoglobulin levels were not routinely collected on patients, which limited our assessment of immune reconstitution after CAR T-cell treatment. Also, the majority of patients returned to their local oncologist for follow-up after the first month after CAR T-cell infusion. While this may have limited data collection, all outside medical records available to the investigators were reviewed. Finally, the COVID-19 pandemic unfolded during the study duration, which may have reduced follow-up and data collection for patients treated after March 2020. Increased respiratory viral testing overall due to COVID-19 may have led to increased detection of microbiologically confirmed viral infections. Yet this difference would not have explained the increased number of viral infections after CD30.CAR T-cell therapy, as patients were assessed during similar time periods after 2020. Notably, the incidence of COVID-19 was comparable between the two cohorts, with six cases (9%) of COVID-19 in CD30.CAR T-cell patients and four cases (8%) in CD19.CAR T-cell patients.

In conclusion, this is the first study to report infections after CD30.CAR T-cell therapy for relapsed/refractory Hodgkin’s lymphoma or peripheral T-cell lymphoma. Identification of reduced ANC prior to CD30.CAR T-cell infusion as a significant factor for infections after cell infusion may serve as a marker for identifying patients at higher risk for infection, although further investigation is needed to replicate these findings in a larger cohort. Given differences in infection risk following treatment, CD30.CAR T-cell treated patients may not benefit as much from antibacterial prophylaxis as much as they might from prophylactic valacyclovir for prevention of HSV and VZV. Additional studies on the infectious risk of novel CAR T-cell therapies are needed with the potential to identify patients at highest risk of developing infections and to tailor antimicrobial prophylaxis after CAR T-cell treatment.

## Data Availability

All data produced in the present study are available upon reasonable request to the authors

## Acknowledgements

The authors would like to acknowledge the staff of the University of North Carolina Chapel Hill and UNC Hospitals, in particular the staff and patients at the Lineberger Cancer Center.

This work was supported in part by the following: F.C. is funded by National Cancer Institute UNC Immunotherapy Training Grant T32 1T32CA28527-01. MM is funded by the Infectious Disease Society of America Grants for Emerging Researchers (G.E.R.M). TMA is funded by the Amy Strelzer Manasevit/Be The Match Foundation Award, a K23 NIAID/NIH Career Development Award (AI163365), and the UNC Center for Gastrointestinal Biology and Disease pilot award. REDCap use was made possible through grant UL1TR002489 from the Clinical and Translational Science Award program of the Division of Research Resources, NIH. J.S.S. is supported by R01 155098 from NHLBI.

## Authorship Contributions

T.M.A., M.S., J.H., J.M., and N.G. conceived of and designed the study with feedback from J.S.S., P.A., G.D., and B.S. who interpreted the data. T.M.A., F.C., M.M., J.S., M.K.S., and B.R. reviewed and collected data from electronic health records and assisted in constructing the tables and figures. Y.X. and H.H. performed the statistical analysis for the project and created the tables and figures. T.M.A. and F.C., wrote the manuscript with significant help from M.M. All authors contributed to the editing of the manuscript and approved the final version.

