## Supplemental Table/Figures for "Infectious Complications Following CD30 Chimeric Antigen Receptor T-Cell Therapy in Adults"

### SUPPLEMENTARY TABLES AND FIGURES

**Table S1.** Demographics and treatment-related complications of patients with and without microbiologically-confirmed infections in one year after anti-CD19 CAR-T therapy.

| | **Infected*** N= 18 | **Not infected** N = 32 | **Total** N = 50 | **p-value** |
|---|---|---|---|---|
| Median age [range] | 48.8 [21.5, 81.0] | 64.4 [27.3, 80.0] | 60.6 [21.5, 81.0] | 0.088 |
| Sex (male), n (%) | 12 (66.7) | 19.0 (59.4) | 31 (62.0) | 0836 |
| Race, n (%) | | | | 0.109 |
| Caucasian | 11 (61.1) | 26 (81.2) | 37 (74.0) | |
| African American | 3 (16.7) | 6 (18.8) | 9 (18.0) | |
| Other | 4 (22.2) | 0 (0.0) | 4 (8.0) | |
| Ethnicity, n (%) | | | | 0.492 |
| Hispanic | 3 (16.7.) | 2 (6.2) | 5 (10.0) | |
| Non-Hispanic | 15 (83.3) | 30 (93.8) | 45 (90.0) | |
| Malignancy, n(%) | | | | **0.040** |
| Acute Lymphocytic Leukemia (ALL) | 8 (44.4) | 4 (12.5) | 12 (24.0) | |
| Diffuse Large B-Cell Lymphoma | 8 (44.4) | 22 (68.8) | 30 (60.0) | |
| Follicular or mantle cell lymphoma | 2 (11.1) | 6 (18.8) | 8 (16.0) | |
| Disease state prior to treatment, n (%) | | | | 0.225 |
| Partial Remission (PR) | 1 (5.6) | 6 (18.8) | 7 (14.0) | |
| Stable Disease (SD) | 1 (5.6) | 4 (12.5) | 5 (10.0) | |
| Progressive Disease (PD) | 15 (83.3) | 22 (68.8) | 37 (74.0) | |
| Very Good Partial Response (VGPR) | 1 (5.6) | 0 (0.0) | 1 (2.0) | |
| KPS score (median [range]) | 85.0 [70.0, 100.0] | 80.0 [60.0, 100.0] | 80.0 [60.0, 100.0] | 0.148 |
| HCT-CI score (median [range]) | 3.0 [2.0, 14.0] | 4.0 [2.0, 8.0] | 4.0 [2.0, 14.0] | 0.203 |
| Prior lines of chemotherapy (median [range]) | 6.0 [2.0, 17.0] | 4.0 [2.0, 10.0] | 4.0 [2.0, 17.0] | **0.017** |
| CAR T lymphodepletion regimen, n (%) | | | | 0.311 |
| Fludarabine/Bendamustine | 1 (5.6) | 4 (12.5) | 5 (10.0) | |
| Fludarabine/Cyclophosphamide | 16 (88.9) | 28 (87.5) | 44 (88.0) | |
| Bendamustine | 1 (5.6) | 0 (0.0) | 1 (2.0) | |
| CAR-T cell product | | | | 0.056 |
| Yescarta | 5 (27.8) | 21 (65.6) | 26 (52.0) | |
| Tecartus | 1 (5.6) | 0 (0.0) | 1 (2.0) | |
| Kymriah | 3 (16.7) | 3 (9.4) | 6 (12.0) | |
| Anti-CD19 trial product | 9 (50.0) | 8 (25.0) | 17 (34.0) | |
| HSCT before CAR T, n (%) | 4 (22.2) | 10 (31.2) | 14 (28.0) | 0.723 |
| HSCT type | | | | 0.574 |
| Allogenic | 3 (16.7) | 5 (15.6) | 8 (16.0) | |

| | | | | |
|---|---|---|---|---|
| Autologous | 1 (5.6) | 5 (15.6) | 6 (12.0) | |
| Median Duration between HSCT and CAR T-cell infusion (Days, median [range]) | 982.0 [529.3, 1699.8] | 1813.5 [1009.8, 2634.3] | 1404.5 [245.0, 9568.0] | 0.396 |
| Auto HSCT within 90 days of CAR T-cell infusion | 2 (11.1) | 2 (6.2) | 4 (8.0) | 0.948 |
| HSCT after CAR T, n (%) | 3 (16.7) | 2 (6.2) | 5 (10.0) | 0.492 |
| Steroids within 1 year following CAR T, n (%) | 12 (66.7) | 9 (28.1) | 21 (42.0) | **0.019** |
| Antimicrobial prophylaxis | | | | |
| Fluoroquinolones | 15 (83.3) | 30 (93.8) | 45 (90.0) | 0.492 |
| Fluconazole | 15 (83.3) | 30 (93.8) | 45 (90.0) | 0.492 |
| Bactrim (trimethoprim-sulfamethoxazole) | 10 (55.6) | 21 (65.6) | 31 (62.0) | 0.689 |
| Valacyclovir | 15 (83.3) | 30 (93.8) | 45 (90.0) | 0.492 |
| Median ANC at Day -30 (median [range]) | 3.9 [0.1, 12.8] | 2.8 [0.1, 19.3] | 3.2 [0.1, 19.3] | 0.266 |
| Median ALC at Day -30 (median [range]) | 1.1 [0.2, 4.0 | 0.5 [0.0, 2.1] | 0.6 [0.0, 4.0] | **0.038** |
| ANC at lymphodepletion (median [range]) | 2.8 [0.1, 14.1] | 3.0 [0.1, 9.1] | 2.8 [0.1, 14.1] | 0.754 |
| ALC at lymphodepletion (median [range]) | 1.1 [0.0, 3.4] | 0.5 [0.0, 1.9] | 0.6 [0.0, 3.4] | **0.008** |
| Total days of neutropenia (median [range]) | 14.0 [0.0, 228.0] | 14.5 [0.0, 161.0] | 14.0 [0.0, 228.0] | 0.895 |
| Total days of lymphopenia (median [range]) | 9.0 [0.0, 86.0] | 9.5 [2.0, 155.0] | 9.5 [0.0, 155.0] | 0.701 |
| Pre-treatment infection within 30 days, n (%) | 3 (16.7) | 4 (12.5) | 7 (14.0) | 1.000 |
| Pre-CART infection organism, n (%) | | | | 0.494 |
| Bacterial | 2 (28.6) | 3 (42.9) | 5 (71.4) | |
| Fungal | 1 (14.3) | 0 (0.0) | 1 (14.3) | |
| Viral | 0 (0.0) | 1 (14.3) | 1 (14.3) | |
| Cytokine-release syndrome (CRS), n (%) | 12 (66.7) | 22 (68.8) | 34 (68.0) | 1 |
| CRS grade, n (%) | | | | 0.131 |
| Grade 1 | 3 (16.7) | 14 (43.8) | 17 (34.0) | |
| Grade 2 | 8 (44.4) | 8 (25.0) | 16 (32.0) | |
| Grade 3 | 1 (5.6) | 0 (0.0) | 1 (2.0) | |
| CRS treatment (Tocilizumab), n (%) | 2 (8.7) | 3 (7.3) | 5 (7.8) | 0.242 |
| Tocilizumab | 9 (50.0) | 9 (28.1) | 18 (36.0) | |
| Tocilizumab + Steroids | 1 (5.6) | 1 (3.1) | 2 (4.0) | |
| Neurotoxicity (ICANS), n (%) | 9 (50.0) | 5 (15.6) | 14 (28.0) | **0.023** |
| ICU Admission (within 30 days of CART infusion, n (%)) | 8 (44.4) | 5 (15.6) | 13 (26.0) | 0.058 |
| Relapse within 1 year after CAR T-cell therapy, n (%) | 10 (55.6) | 18 (56.2) | 28 (56.0) | 1 |
| Cause of Death | | | | **0.004** |
| Infection-related | 7 (63.6) | 0 (0.0) | 7 (14.0) | |
| Relapse-related | 4 (36.4) | 13 (86.7) | 17 (34.0) | |
| Unknown | 0 (0.0) | 1 (6.7) | 1 (2.0) | |
| Other | 0 (0.0) | 1 (6.7) | 1 (2.0) | |

| | | | | |
|---|---|---|---|---|
| Mortality within 30 days of CAR T, n (%) | 0 (0.0) | 0 (0.0) | 0 (0.0) | NA |

Footnote:
*33 total infections in 18 patients during the follow-up period.
[1] KPS, Karnofsky Performance Status
[2] HCT-CI, Hematopoietic Stem Cell Transplant-specific Comorbidity Index
[3] HSCT, Hematopoietic Stem Cell Transplant
[4] Steroids (30 days prior to CAR T through 1 year after CAR T) include dexamethasone, prednisone, and methylprednisolone
[5] ANC, absolute neutrophil count
[6] ALC, absolute lymphocyte count
[7] Neutropenia defined as ANC <500 cells/$mm^3$
[8] Lymphopenia defined as ALC <200 cells/$mm^3$
[9] CRS was only treated with Tocilizumab in this cohort

**Figure S1**. **Number of infections after CAR T-cell therapy.** Number of infections including bacterial and viral etiologies (no fungal infections were seen) during the first 1 year following CAR T-cell therapy, broken into infection periods (0-28, 29-90, and 91-365 days) and compared between anti-CD30 CAR-T therapy (A) and anti-CD19 CAR-T therapy (B). Data shown is censored for relapse.

A.

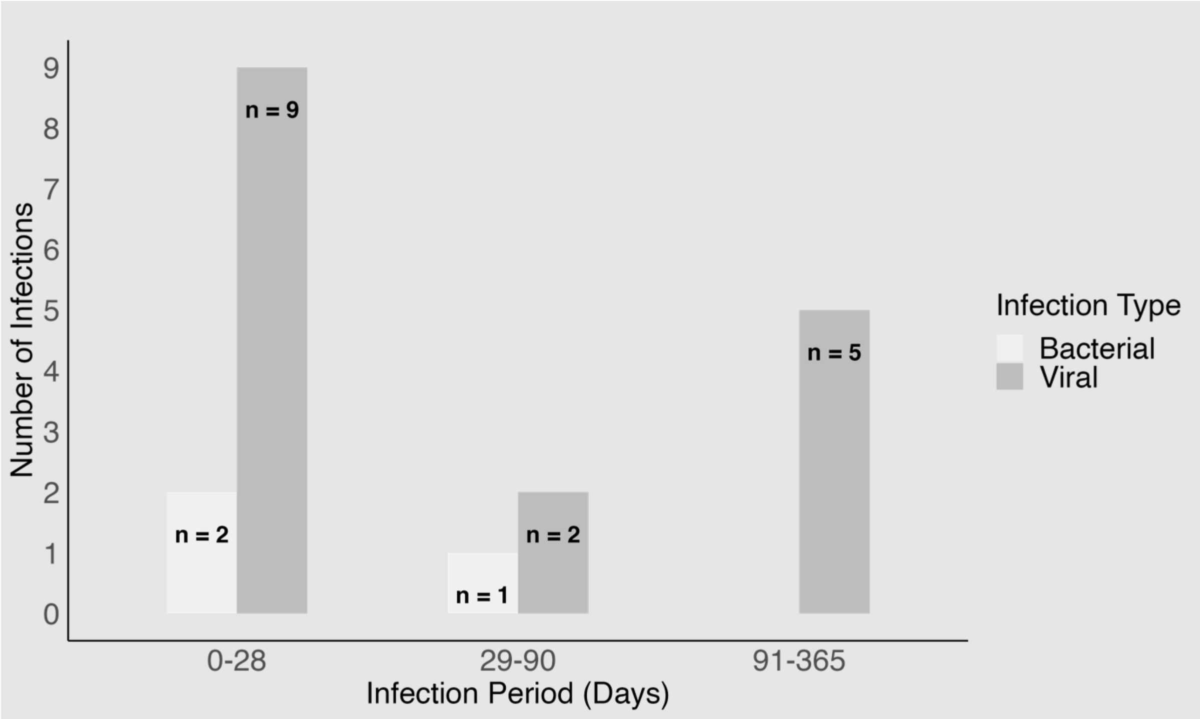


B.

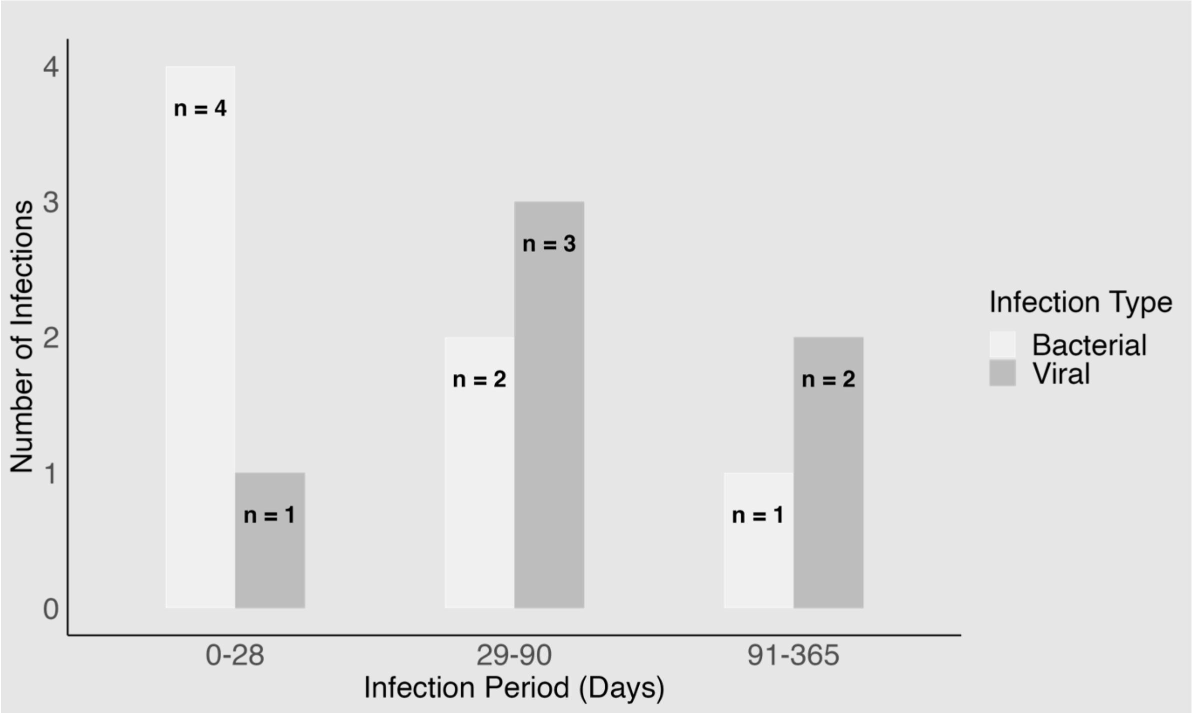

**Figure S2. Severity of infections after CAR T-cell therapy.** Number of infections including bacterial and viral etiologies (no fungal infections were seen) of different severities occurring during the first 1 year following CAR T-cell therapy, broken into infection periods (0-28, 29-90, and 91-365 days), and compared between anti-CD30 CAR-T therapy (A) and anti-CD19 CAR-T therapy (B). Data shown is censored for relapse.

A.

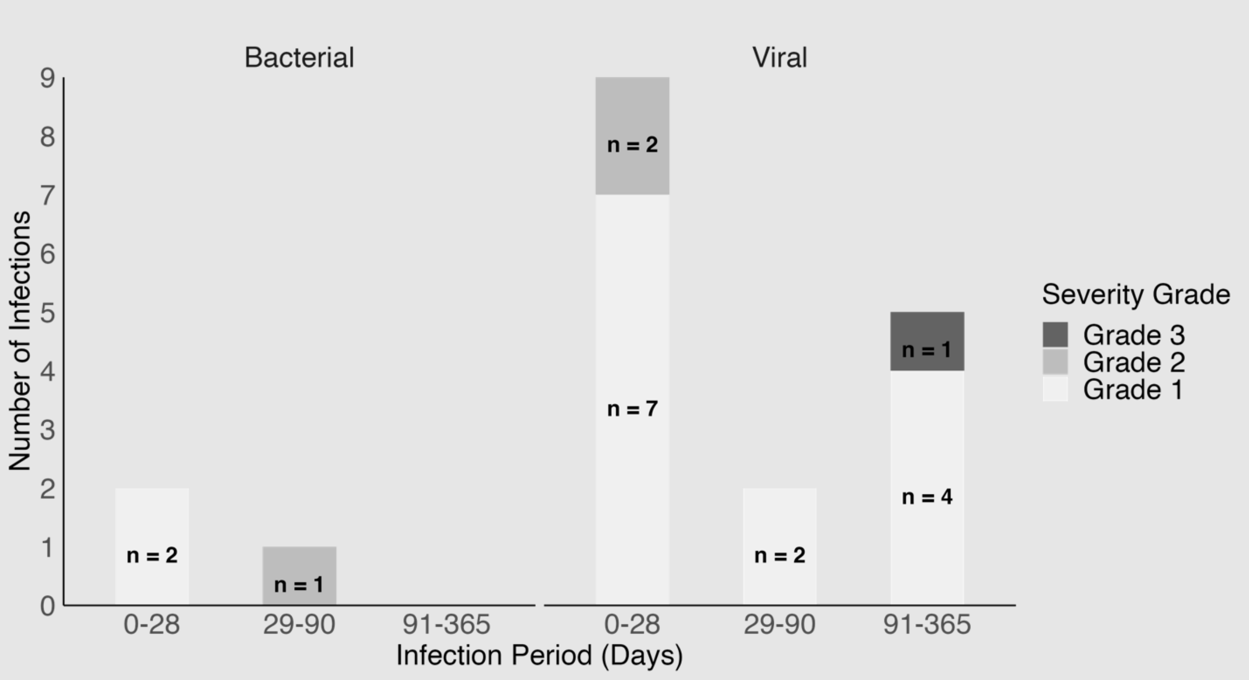


B.

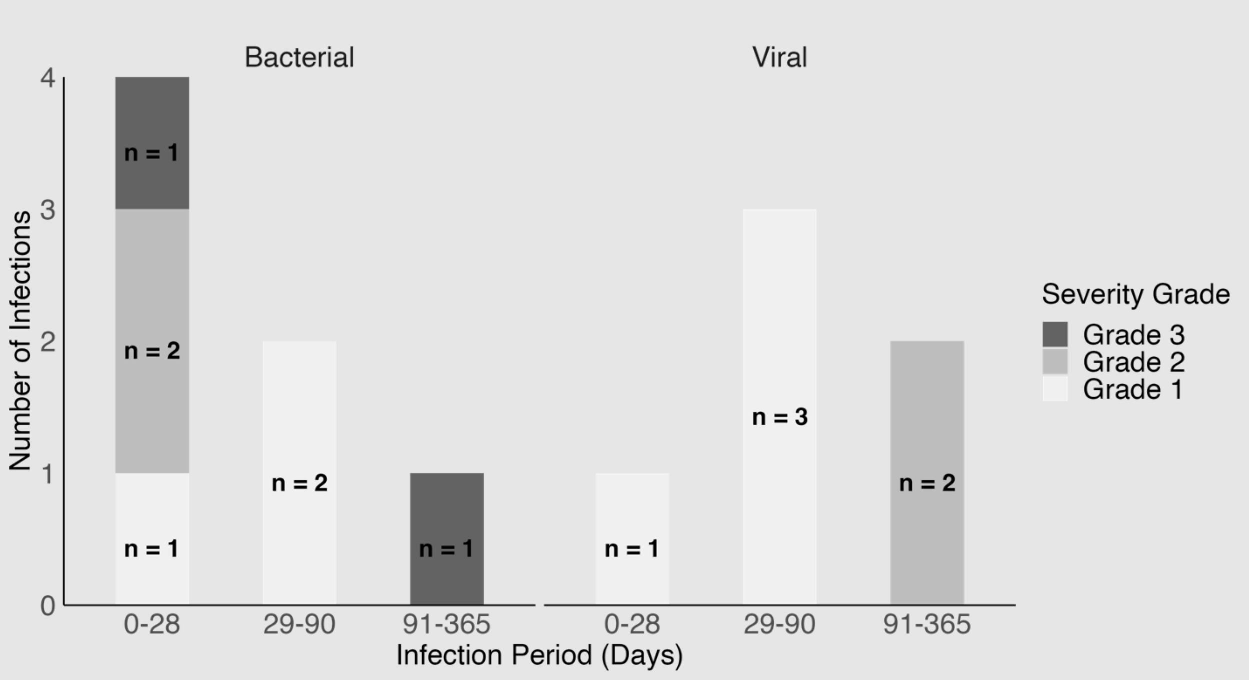

**Figure S3**. **Immune recovery after CD19 CAR T-cell therapy demonstrates rare differences between infected and uninfected patients within 1 year after infusion**. Absolute neutrophil counts (ANC, Figure 4A) and absolute lymphocyte counts (ALC; Figure 4B) are demonstrated at timepoints relative to the day of CAR-T infusion. Comparisons between infected and uninfected patients were performed using the Wilcoxon rank-sum test, with p-value <0.05 being given one star at the top of the plot.

A.

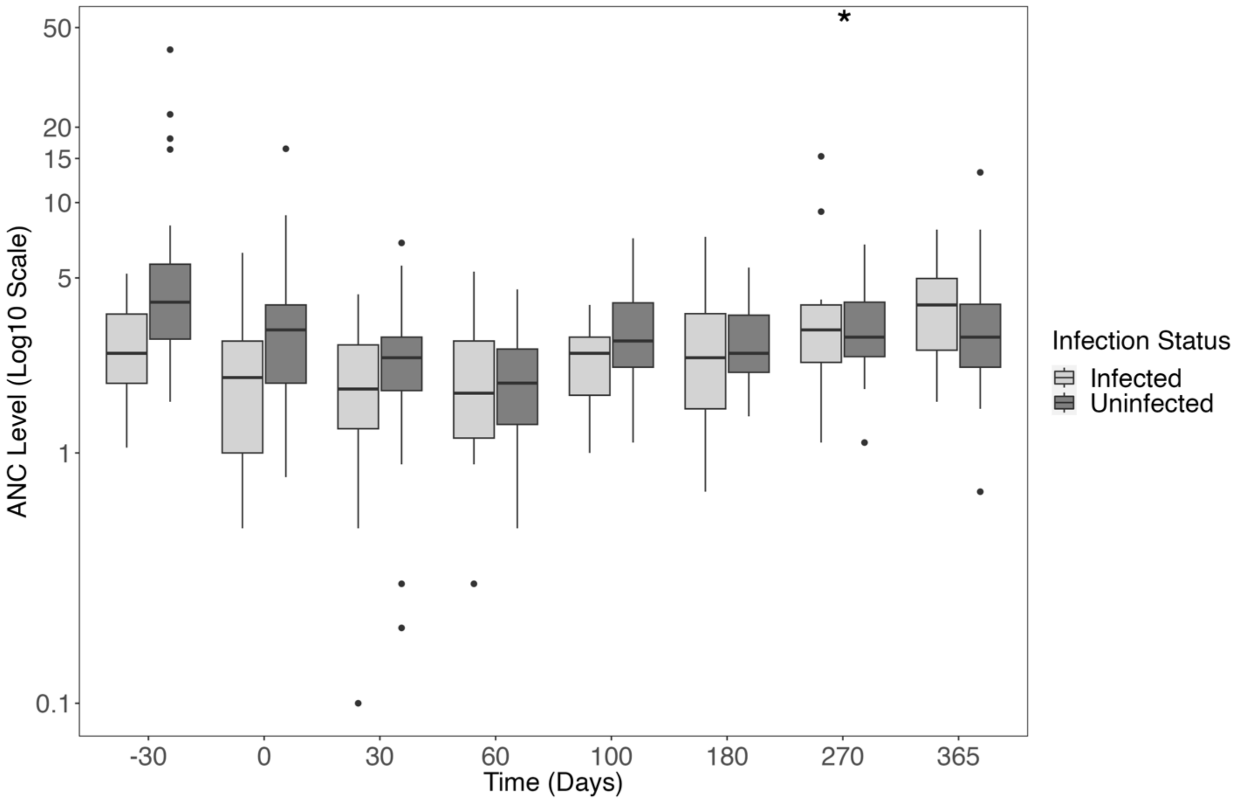

B.

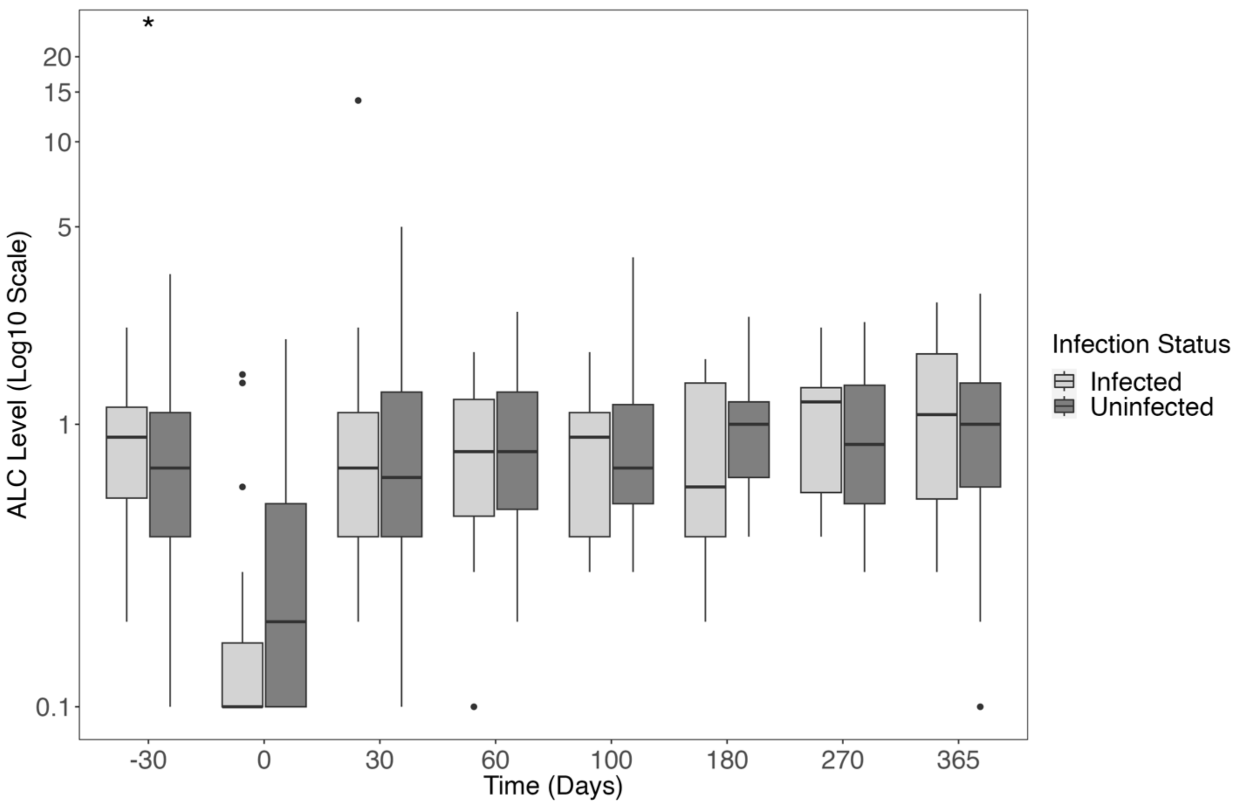

*
20
15
10
5
1
0.1
ALC Level (Log10 Scale)
-30
0
30
60
100
180
270
365
Time (Days)
Infection Status
Infected
Uninfected
